# The Effect of Headgear Use on Concussion Injury Rates in High School Lacrosse

**DOI:** 10.1101/2021.10.06.21264026

**Authors:** Daniel C. Herman, Shane V. Caswell, Patricia M. Kelshaw, Heather K. Vincent, Andrew E. Lincoln

## Abstract

**Objectives:** The use of headgear is a controversial issue in girls’ lacrosse. We compared concussion rates among high school lacrosse players wearing versus not wearing lacrosse headgear.

**Methods:** Study participants included a sample of convenience of high schools with girls’ lacrosse from across the United States. Certified athletic trainers reported athlete exposure and injury data via the National Athletic Treatment, Injury and Outcomes Network during the 2019 through 2021 seasons. The Headgear cohort was inclusive of high schools from the state of Florida, which mandate the use of ASTM standard F3137 headgear, while the Non-Headgear cohort was inclusive of the remaining states, none of which have headgear mandates. Incidence Rate Ratios (IRR) and 95% Confidence Intervals (CI) were calculated. IRRs with corresponding CIs that excluded 1.00 were deemed statistically significant.

**Results:** 141 concussions (Headgear: 25; Non-Headgear: 116) and 357,225 Athlete Exposures (AE) were reported (Headgear: 91,074AE; Non-Headgear: 266,151AE) across all games and practices. Overall, the concussion injury rate per 1000AE was significantly higher in the Non-Headgear cohort (0.44) than the Headgear Cohort (0.27) (IRR=1.59, 95% CI:1.03 - 2.45). The IRR was significantly higher for the Non-Headgear cohort during games (1.74, 95% CI: 1.00, 3.02) but not for practices (1.42, 95% CI: 0.71, 2.83).

**Conclusions:** These findings indicate that concussion rates among high school girls’ lacrosse players not wearing headgear were 59% higher than those wearing headgear. These data support the use of protective headgear to reduce the risk of concussion among high school female lacrosse athletes.

**Summary:** *What are the new findings:*

- The use of lacrosse headgear meeting the ASTM F3137 standard was associated with a lower risk of experiencing a concussion injury among high school girls’ lacrosse players. How might these findings impact clinical practice in the future?

- Lacrosse headgear may be warranted for use for concussion risk mitigation among high school girls’ lacrosse players.
- Lacrosse headgear may be considered for concussion risk mitigation at other levels of play such as the youth or collegiate levels; further study is warranted.

## Introduction

Girls’ lacrosse continues to be the fastest growing high school sport in the United States (US).[1] Prior to the COVID-19 pandemic, high school girls’ lacrosse participation grew by 53.63% over the past decade in the US (2008-2009: n=64,929; 2018-2019: n=99,750).[1] Girls’ lacrosse is a noncontact sport; however incidental concussions[2-5] and head impacts[6-8] associated with lacrosse game-play are common. A recent epidemiological study, observed across five years (2008-09 through 2013-14) of competitive high school girls’ lacrosse, demonstrated that head/face injuries accounted for the most common game-related injuries (0.92/1000 Athlete Exposures [AE]), the majority of which were concussion (0.83/1000AE).[5] Collectively, stick contact is the leading mechanism of both head impacts[7] and subsequent concussions[5] in high school girls’ lacrosse.

Due to the non-contact rules for girls’ lacrosse, mandated protective equipment is limited to mouthguards and eyewear.[9 10] However, in response to the growing concerns regarding the mechanisms of concussion in girls’ lacrosse from incidental contact, rules allowing for the use of soft-shell headgear have been adopted as of January 1, 2017.[9] Specifically, girls’ lacrosse headgear must meet the ASTM International F3137 performance standard.[11] As stated in the standard, the headgear was designed “*…to address the forces of some incidental stick and ball to headgear impacts to non-goaltending field players*.*”*[1] However, headgear effectiveness at mitigating concussion risk remains unclear.[6 12] Despite this lack of clarity, in 2018 the Florida High School Athletic Association mandated the use of headgear meeting the ASTM standard headgear for lacrosse participation.

Substantial debate exists among the lacrosse community regarding implementing headgear in a noncontact sport that often exhibits incidental contact-related injury.[13-16] Competing arguments have pitted the potential benefits of protective headgear (i.e. decreasing severity of impacts could thus decrease the risk of injury) versus the potential for increased aggression and related injury due to risk compensation (i.e. the Peltzman Effect).[17] Until now, there has been insufficient evidence regarding the effectiveness of lacrosse headgear for reducing the risk of concussion among girls’ lacrosse players to support either argument. Preliminary evidence at the high school level of girls’ lacrosse suggests that headgear may be associated with reduced head impact magnitudes,[6] may not be associated with risk compensatory behaviors,[18] and may be associated with lower rates of concussion injuries during game-play.[19] Therefore, in an effort to better evaluate the implementation of headgear in high school girls’ lacrosse, we compared concussion rates among high school girls’ lacrosse players in the United States wearing headgear versus without headgear. We hypothesized that the rates of concussion among those players wearing headgear would not be different compared to those players not wearing headgear.

## Methods

Our approach utilized a quasi-experimental comparison that leveraged the mandatory policy of headgear use in the state of Florida, and compared outcomes to non-headgear-mandating states across the United States (US). This approach utilized an existing national high school injury registry: the High School National Athletic Treatment, Injury and Outcomes Network (NATION). NATION is an athletic trainer-driven injury reporting registry which utilizes a set of common data elements. It is administered by the Datalys Center for Sports Injury Research and Prevention (Indianapolis, IN) and has numerous publications stemming from its use.[20] Patients or the public were not involved in the design, or conduct, or reporting, or dissemination plans of our research

### Recruitment

In order to obtain a high number of player activity exposures, high school athletic trainers were recruited to report data for high school girls’ lacrosse to NATION. Athletic trainers (ATs) were incentivised with a $150 payment for each full season of reporting to NATION. ATs were recruited using publicly available contact information, outreach in collaboration with the National Athletic Trainers Association and the Athletic Training Locations and Services Project, and word-of-mouth. ATs were eligible to report data for the study if: (i) their high school offered a school sponsored girls’ lacrosse team, (ii) the AT provided regular onsite care and athletic training services to the athletes, and (iii) the status of a headgear mandate at the local level (i.e., Florida teams must implement headgear, versus teams from non-Florida states could not mandate headgear).

### Data Reporting

ATs who agreed to participate were referred to Datalys Center, Inc, which provided training on account management and use of the system to report athlete exposure and injury data. A detailed description of the NATION injury-surveillance methods has been published.[21] In brief, ATs who participate in NATION injury-surveillance efforts collected and entered injury and exposure data into a certified electronic medical record that enabled the exporting of data to NATION.[21] Deidentified exposure and injury data were then extracted from these records and checked for errors by trained, experienced NATION data quality-control staff.[21] The NATION injury-surveillance registry has been approved by the Western Institutional Review Board (Puyallup, WA), and the current investigation was approved by the Institutional Review Board (IRB #201802880) at the University of Florida.

Reported concussions were operationally defined as injuries that occurred as a result of participation in a girls’ high school lacrosse game or practice and were diagnosed by an AT, physician, or other health care professional.[21] Mechanisms of concussion were categorized as player contact, surface contact, contact with equipment, or other/unknown. Time loss for the concussion injuries was defined as the number of days from the injury to return to play. An athlete-exposure (AE) was defined as a single athlete participating in one high school-sanctioned practice or game, regardless of duration, in which the athlete was exposed to the risk of injury.[21] A game exposure required that the athlete participate in the game event to be considered exposed (i.e., athletes on the sideline were not included).[21] Per USA Lacrosse guidelines, individual players from the No Headgear cohort (i.e., non-Florida states) were not restricted from the using headgear during lacrosse participation, and the de-identification process used by High School NATION precluded segregation of injuries and AEs from players using headgear in the No Headgear cohort.

### Power Analysis

A power analysis was performed using publicly available data from the High School Reporting Information Online data registry.[22] Using data from high school girls’ lacrosse from 2014 through 2017, an estimate of 1.4 total concussions (from games and practices) per 1000 game exposures was used to approximate a baseline non-helmeted player concussion injury rate. A Poisson-distributed model with 80% power and 5% type 1 error rate and a 1:2 cohort ratio was then used to obtain an estimate of 24,509 game exposures in Florida and 49,018 exposures outside of Florida in order to resolve a difference of one concussion per 1000 game exposures. Using an estimate of approximately 375 game exposures per high school (i.e., 25 players over 15 games), approximately 65 school-seasons in Florida and 130 school-seasons outside of Florida would be need to provide appropriate power for the study.

### Data Analysis

Concussion incidence rates (IRs) were calculated as the number (i.e., frequency) of documented concussions divided by the athletic-exposures for games or practices where players had an opportunity to experience a concussion multiplied by 1000. Concussions experienced by goalkeepers were excluded from the analyses. Incident rate ratios (IRRs) were calculated as the ratio of the incident rate for the No Headgear cohort divided by that for the Headgear cohort. 95% confidence intervals (95% CI) were calculated for both rates and rate ratios using standard techniques. Exploratory analyses were conducted of incidence rates based on mechanism of injury. IRRs with corresponding CIs that excluded 1.00 were deemed statistically significant.

## Results

A combined 76 school-seasons of data from high schools in the state of Florida and 166 school-seasons of data from high schools outside of the state of Florida were reported during the 2019 and 2021 seasons (See Table 1). Twenty partial school-seasons of data from high schools in the state of Florida and 27 partial school-seasons of data from high schools outside of the state of Florida were reported during the 2020 season prior to cancellation due to the coronavirus pandemic. Table 1 includes the AEs reported during each of these seasons.

**Table 1.** Athlete Exposures

|  | No Headgear | Headgear | Overall |
| --- | --- | --- | --- |
| Team-Seasons |  |  |  |
| 2018-2019 | 48 | 26 | 74 |
| 2019-2020* | 27 | 20 | 47 |
| 2020-2021 | 118 | 50 | 168 |
| Total | 193 | 96 | 289 |
| Athlete Exposures |  |  |  |
| 2018-2019 | 89,504 | 26,321 | 115,825 |
| 2019-2020* | 6,656 | 6,246 | 12,902 |
| 2020-2021 | 169,991 | 58,507 | 228,498 |
| Total Games | 73,125 | 25,733 | 98,858 |
| Total Practices | 193,026 | 65,341 | 258,367 |
| Total Games and Practices | 266,151 | 91,074 | 357,225 |

\*Season shortened as a result of the COVID-19 pandemic.

Over the three seasons there were a total of 141 documented concussions that occurred over 357,225 AEs, resulting in an overall rate of 0.39 concussions per 1000 AE (95% CI: 0.33, 0.46) (See Table 2). The Headgear cohort experienced 25 concussions (17.7%, IR=0.27 per 1000 AE, 95% CI: 0.17, 0.38) while the No Headgear cohort had 116 concussions (82.3%, IR=0.27 per 1000 AE, 95% CI: 0.36, 0.52). The IRs during games were dramatically higher for both the Headgear (0.58, 95% CI: 0.29, 0.88) and No Headgear (1.01, 95% CI: 0.78, 1.24) cohorts than during practices. The IRRs were significantly higher for the No Headgear cohort during games (1.74, 95% CI: 1.00, 3.02) and overall (1.59, 95% CI: 1.03, 2.45), but not for practices (1.42, 95% CI: 0.71, 2.83).

**Table 2.**
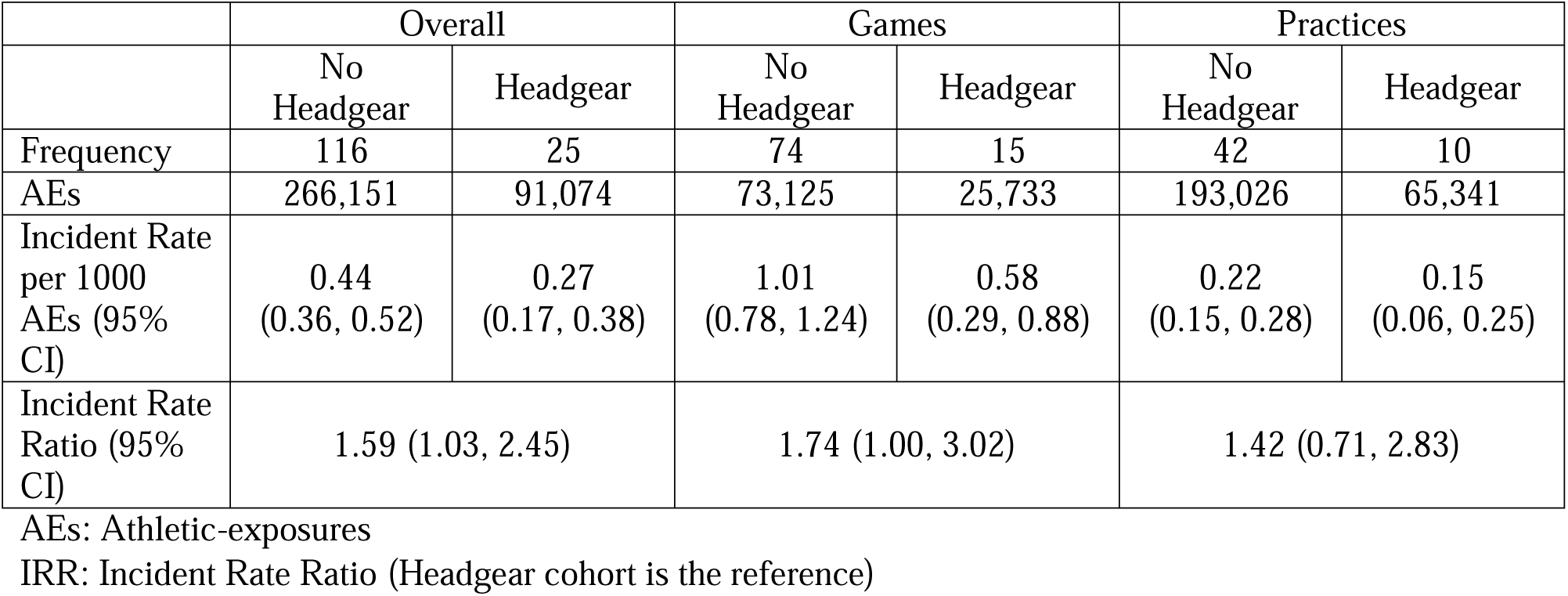
Concussion incidence among girls’ high school lacrosse players wearing headgear vs. not wearing headgear by game type

In terms of mechanism of injury, contact with stick/ball resulted in the majority of concussions (n=74, 52.5%), followed by contact with player (n=38, 27.0%) and contact with ground (n=26, 18.4%) (See Table 3). The IRRs were not significantly higher among the No Headgear cohort for player contact with equipment (1.77, 95% CI: 0.95, 3.28), player (2.26, 95% CI: 0.88, 5.79), or ground (0.93, 95% CI: 0.39, 2.21).

**Table 3.** Concussion incidence among girls’ high school lacrosse players wearing headgear vs. not wearing headgear by mechanism of concussion

|  | Equipment |  | Player |  | Ground |  |
| --- | --- | --- | --- | --- | --- | --- |
|  | No Headgear | Headgear | No Headgear | Headgear | No Headgear | Headgear |
| Frequency | 62 | 12 | 33 | 5 | 19 | 7 |
| AEs | 266,151 | 91,074 | 266,151 | 91,074 | 266,151 | 91,074 |
| Incident Rate per 1000 AEs (95% CI) | 0.23<br>(0.17, 0.29) | 0.13<br>(0.06, 0.21) | 0.12<br>(0.08, 0.17) | 0.05<br>(0.01, 0.10) | 0.22<br>(0.15, 0.28) | 0.08<br>(0.02, 0.13) |
| Incident Rate Ratio (95% CI) | 1.77 (0.95, 3.28) |  | 2.26 (0.88, 5.79) |  | 0.93 (0.39, 2.21) |  |

AEs: Athletic-exposures
IRR: Incident Rate Ratio (Headgear cohort is the reference)
“Other” mechanism (n=3) not included

## Discussion

Our findings provide much needed information about concussion rates among high school girls’ lacrosse players in Florida, which mandates lacrosse headgear, compared to those states having no headgear mandate. We observed that girls’ participating in states not mandating lacrosse headgear had a 59% greater overall incidence of concussion than those required to wear headgear. Moreover, a 74% greater incidence of concussion was observed during game play in states not mandating headgear. However, headgear use was not associated with any significant differences in concussion incidence during practice. Collectively, these findings suggest that use of lacrosse headgear is associated with a lower incidence of concussion incidence in girls’ high school lacrosse.

### Comparison to Previous Investigations

Our findings are consistent with a previous smaller investigation that studied this issue regionally.[19] Baron et. al. followed girls’ high school lacrosse athletes from the Public Schools Athletic League of New York City, which has a headgear mandate in place. The authors compared concussion rates among their cohort from the 2017 and 2018 seasons to that of the High School RIO nationwide injury registry from 2009-2016. The use of headgear was associated with a lower overall concussion rates (0.089 per 1000AE) compared to the nationwide cohort (0.375 per 1000AE).

While the concussion rates of the headgear cohort were low compared to the headgear cohort in the present investigation, the study by Baron et. al. was limited by the relatively small number of athlete exposures (22,397AE) among the headgear cohort. In contrast, our study leveraged over four times the headgear cohort AE of this prior study. Furthermore, we used a contemporaneous cohort without headgear for comparison, as opposed to Baron et. al. which used historical data from a previous period with a much wider range of seasons. Additionally, game exposures in our study consisted of all participants playing with headgear or all participants playing without headgear; conversely, the game exposures in the headgear group for Baron et. al. consisted of games with one or both teams wearing headgear. Finally, all of the data recorded in this study were derived from the one data collection registry with one data collection technique rather than a combination of methodologies. These advantages allow for a more robust interpretation of the associations of concussion incidence with headgear use in girls’ lacrosse.

### Mechanisms of Injury

Although not the primary purpose of the study, the relationship between headgear use, concussion and mechanisms of injury were explored. The findings show there was a reduction in the rate of concussion by nearly half with headgear in situations where equipment (i.e. ball or stick) versus player were involved. While this was not statistically significant, this is a compelling avenue for additional research. Prior research suggests the most common mechanism of concussion in girl’s lacrosse is via equipment versus player contact.[5] The magnitude of impacts associated with this mechanism of injury have the potential to be particularly high, with linear and rotational acceleration magnitudes from ball and stick impacts second only to falls.[23] Thus, any reduction in concussion risk from this mechanism is clinically important. Previous investigations support the ability of headgear meeting the ASTM F3137 standard in reducing impact magnitudes. Laboratory research using lacrosse headgear meeting the ASTM F3137 standard report a reduction in both linear and rotational accelerations.[24] Similarly, a recent study investigated the effect of headgear versus no headgear conditions on peak linear accelerations and peak rotational velocities during actual game play on high school girls lacrosse players instrumented with wearable sensors.[6] The headgear condition resulted in slight reductions to the mean impact magnitudes experienced by the players.[6] Our findings may lend support to the notion that the headgear are effective at mitigating such impacts that may result in concussion among this cohort; however, further investigation is needed using study designs that are appropriately powered to assess concussion incidence based on mechanism of injury.

Similarly, there was a non-significant reduction in the incidence of concussion by a player versus player mechanism of injury by over half with headgear use. We are unable to discern a rationale for a potential protective effect of lacrosse headgear for concussions resulting from player contact mechanisms; however, our data may provide insight into the potential role of risk compensation, also known as the Peltzman Effect.[17] This postulates that individuals may act with less caution when they have a greater level of protection; or, when applied to gameplay scenarios such as lacrosse, players may act with greater aggression towards an opponent that they feel has a greater level of protection. If the headgear was protective but significant risk compensation also occurred, players in the headgear cohort may have experienced a reduction in equipment vs player concussions but a greater rate of player versus player or player versus ground concussions. However, this was not observed, which speaks against any effect of risk compensation on the study results. Collectively, our findings along with prior observations of perceptions of headgear use,[18] may dispel some concerns regarding compensatory aggressive behaviors subsequent to headgear use in girls’ lacrosse. Again, given the fact that our study was not adequately powered to make comparisons based on mechanism of injury, this possibility should be treated with caution.

### Limitations

The primary limitation of this study is the lack of randomisation regarding the use of protective headgear. The use of players at high schools in the state of Florida for the headgear cohort was necessary owing to the fact that Florida is the only state in the United States with a protective headgear mandate for girls’ lacrosse. Regardless, this design may introduce potential confounding elements that would bias the results. Possible confounders may be regional differences in game play and officiating between the state of Florida and the remainder of the United State.

It was assumed that all players in the No Headgear cohort in fact did not use headgear; however, as noted previously, players were not restricted from using headgear and the nature of the reporting system precluded segregation of data from such players in the No Headgear cohort. Anecdotally, the use of headgear among states outside of Florida and in areas without a local helmet mandate is exceedingly low, and thus we feel the contribution of injury and AE data from athletes using headgear in the No Headgear cohort to be trivial. Furthermore, given the demonstrated association of headgear use with a reduced risk of concussion, any contributions from such athletes in the No Headgear cohort would serve to strengthen the findings rather than diminish the relationship.

However, this assumption does pose another limitation to the current study. The comparison cohorts included games in which all players used headgear versus when all players did not use headgear; as such we are unable to make conclusions regarding games featuring players in games with mixed headgear status (e.g. one team using headgear while the other team does not). It is possible that any contribution from risk compensation may be different under such circumstances, although the prior biomechanical study by Caswell et. al., which was conducted using such an environment, suggests otherwise.[6]

### Conclusion and Future Directions

The results of this study are highly encouraging for athlete safety in high school girls’ lacrosse. The data indicate that the use of protective headgear is associated with significantly lower risk of experiencing a concussion injury; as such, we feel that this supports the use of the ASTM F3137 standard headgear for the sport in this age cohort.

It is possible that protective headgear may have similar effects in different populations of girls’ lacrosse. These may include athletes at the collegiate level or higher, as well as athletes at the developmental or youth levels. A measure of caution is necessary as significant differences in game play, officiating performance, body control, skill development, and neck strength are likely to be present at these difference levels, which may influence the association between headgear and concussion risk. In light of the current results, additional investigation is warranted for these populations.

These findings also suggest that headgear may be protective in other non-contact sports with moderate-high rates of concussion that do not mandate protective equipment. Formal evaluation studies in such sports would be needed to confirm any protective effects and detect any potential unintended consequences.

## Data Availability

Data may be available upon request.

